# Secondary transmission of SARS-CoV-2 in educational settings in Northern Italy from September 2020 to April 2021: a population-based study

**DOI:** 10.1101/2021.09.03.21263061

**Authors:** Olivera Djuric, Elisabetta Larosa, Mariateresa Cassinadri, Silvia Cilloni, Eufemia Bisaccia, Davide Pepe, Massimo Vicentini, Francesco Venturelli, Laura Bonvicini, Paolo Giorgi Rossi, Patrizio Pezzotti, Alberto Mateo Urdiales, Emanuela Bedeschi, the Reggio Emilia Covid-19 Working Group

## Abstract

**Background:** We aimed to quantify the risk of transmission of SARS-CoV-2 in the school setting by type of school, characteristics of the index case and calendar period in the Reggio Emilia province (RE), Italy. The secondary aim was to estimate the promptness of contact tracing.

**Methods:** A population-based analysis of surveillance data of all COVID-19 cases occurring in RE, Italy, from September 1, 2020, to April 4th, 2021, for which a school contact and/or exposure was suspected. Indicator of the delay in contact tracing was computed as the time elapsed since positivity of the index case and the date on which the swab for classmates was scheduled (or most were scheduled).

**Results:** Overall, 30,184 and 13,608 contacts among classmates and teachers/staff, respectively, were identified and received recommendation for testing; 43,214 (98.7%) performed the test. Secondary transmission occurred in about 40% of the investigated classes, and the overall secondary case attack rate was 4%, slightly higher when the index case was a teacher, but with almost no differences by type of school and stable during the study period. Promptness of contact tracing increased during the study period, reducing the time from index case identification and testing of contacts from 7 to 3 days, as well the ability to identify possible source of infection in the index case.

**Conclusions:** Despite the spread of the Alpha variant during the study period in RE, the secondary case attack rate remained stable from school reopening in September 2020 until the beginning of April 2021.

## Background

Since the first case of COVID-19 was described at the end of 2019, there have been almost 249 million cases and 5 million deaths reported globally up to October 2021.^1^ In the period prior to the implementation of mass vaccination programmes, social distancing and movement-restriction measures were the main measures undertaken to prevent virus transmission. Closure of educational institutions was one of the preventive measures considered and often adopted during the pandemic. This was due to the concern about potential school-to-home transmissions of the virus from students to more susceptible family members, although the overall risk of severe COVID-19 in children and young people was shown to be very small.^2^

The role of school contacts in the spread of the virus as well the effectiveness of school closures in controlling the epidemic have been debated, and inconsistent results have emerged.^3,4^ Modelling studies have provided scenarios with a small impact of school closure on virus transmission under the conditions hypothesized by the authors.^5,6,7^ A recent systematic review of empiric studies^8^ showed heterogenous results ranging from no effect to an important reduction in virus transmission in the community, with studies with lower risk of bias showing a small effect.^9,10^ Finally, three large studies from the US, the UK and Italy suggest a limited role of school transmission in determining mortality in the community.^11-13^ It is worth noting that school opening may impact on virus transmission because of the increase in public transport-related contacts and other non-school activities, not only through contacts occurring in the classroom.

In our first study conducted between 1 September and 15 October 2020, non-negligible secondary attack rates were detected, especially in secondary schools. After this first report, several changes occurred that could have influenced the risk of transmission and its control in schools.^14^ Starting from 27 November 2020, the local health authority improved contact tracing protocols and introduced immediate molecular tests for all contacts, whether symptomatic or asymptomatic, at the beginning of quarantine, with the aim of identifying all possible sources of infection in asymptomatic contacts, i.e., backward tracing (figure 1). Policies to reduce crowding especially in high schools were introduced (reducing in-class time by 25% to 50%) as were several short closures in the periods of highest incidence (figure 2). Finally, at the end of December 2020, the Alpha variant started to circulate in our province, becoming predominant in February 2021.^15^

**Figure 1.**
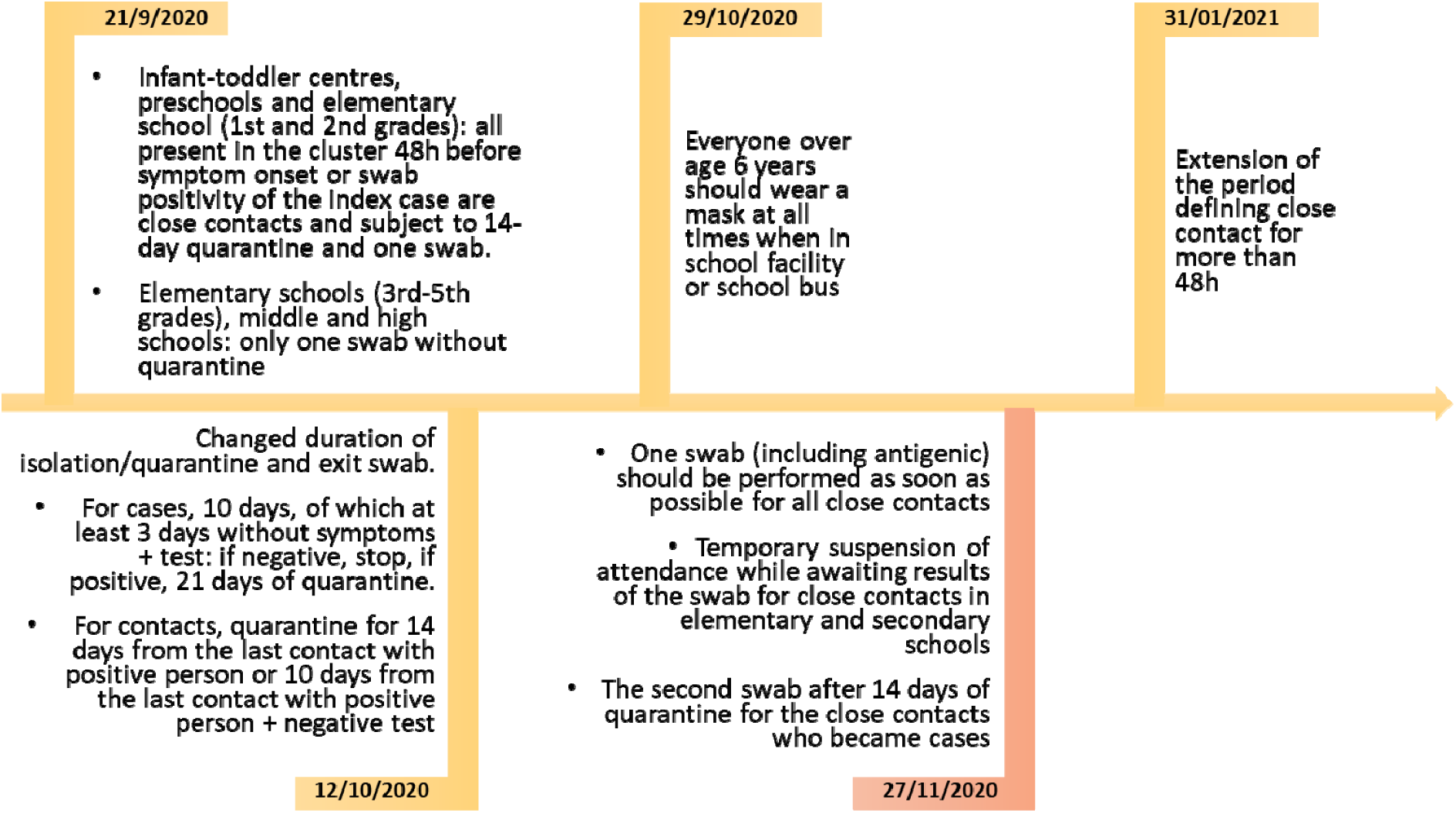
Timeline of the major changes in regulations and recommendations for management of COVID-19 cases in educational settings in Reggio Emilia province

**Figure 2.**
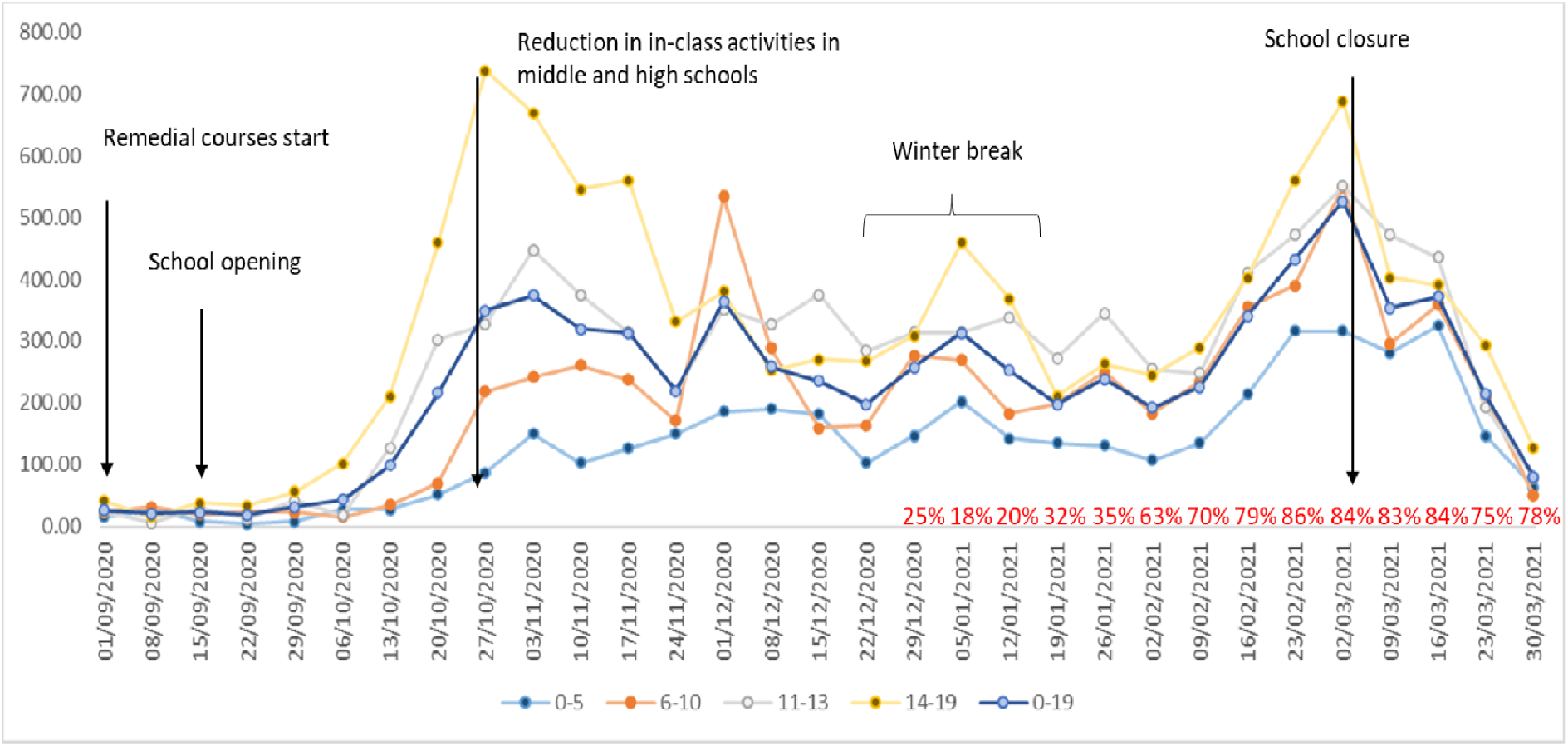
Weekly notification rates of new COVID-19 cases per 100,000 inhabitants aged 0-19 years, by age class, Reggio Emilia province, 1 September 2020 – 31 March 2021. In the graph the main changes in school opening and closure are also reported, and (in red) the proportion of Alpha variant among sequenced cases as reported by the Italian National Institute of Health.

In this study we present the results of comprehensive epidemiological investigations conducted from school reopening in September 2020 until the end of March 2021. Our aim was to quantify the risk of infection transmission in the school setting by type of school, characteristics of the index case and calendar period in the Reggio Emilia province by analysing surveillance data about all cases who had contacts in the school setting. The second aim was to estimate the promptness of contact tracing.

## Methods

### Setting

There are approximately 95,000 inhabitants form 6 months-old to 19-year-old that can attend infant-toddler centres (ages 0-3 years), preschools (ages 3-5), primary schools (ages 6-10), middle schools (ages 11-13) and high schools (ages 14-19), and about 12,000 teachers/ school staff members in Reggio Emilia province (531,751 inhabitants, Emilia Romagna, Northern Italy). During the study period, there were two peaks of infections: in November 2020 and in February/March 2021 (figure 2). After the school reopening on 1 September 2020 for remedial courses and on 15 September 2020 for the regular school year, in-class learning was in place until 26 October 2020, when most high schools moved to remote learning for at least 75% of scheduled lessons. In addition, because of the to the high circulation of the virus, the Christmas school holidays were extended to the second week of January, i.e., from 20 December to 11/15 January. Another lockdown led to the closing of schools on 3 March 2021 (figure 2). Only infant-toddler centres and preschools, schools that requires laboratory work and schools with pupils with disabilities or special needs continued in-class didactic activities.

### Design

An analysis of population-based surveillance data was conducted including all consecutive positive cases confirmed with RT-PCR for SARS-COV-2 infection between 1 September 2020 and 4 April 2021 in Reggio Emilia province that led to an epidemiological investigation among children and adolescents (0–19 years) who may have had exposure or contact with positive cases at school. Given that reporting positivity of even one child led to a field investigation, contact tracing and, in some cases, quarantine measures, all cases were included, regardless of the presence of an outbreak or not. We excluded cases that occurred among children not attending schools in the period investigated for contact tracing, i.e., in the period starting 48 h before symptom onset, and for asymptomatic cases, 48 h before diagnosis or 48 h after contact with a certain case, whichever occurred first.

### Control measures

When a case occurred in primary and secondary schools, in-class activities were usually suspended, but quarantine was not mandatory if all physical distancing measures were respected (sufficient distance in the classroom, physical distance during all activities and utilization of face masks). All students and school personnel were asked to perform a swab, with an appointment scheduled as early as possible and communicated to the parents by means of a text message. If at least one secondary case was found, quarantine was mandatory for all students in that class. For all close contacts identified during the epidemiological investigation, a 10-day quarantine and an exit negative test were required. In the infant-toddler and preschool settings, both the teachers and children were considered close contacts if they had had contact with a case at school 48h before symptom onset or a positive swab, in which case they were subjected to quarantine. In some cases, based on the surveillance team’s evaluation, the entire school was closed. Teachers were never quarantined for school contacts, except if they taught in an infant-toddler centre or preschool. Other control measures have been described more in detail elsewhere^14^; the major changes occurring during the study period are described in figure 1.

### Outcome definition and variables of interest

The main outcome was the secondary transmission rate. Overall attack rates were calculated by dividing the number of cases by the population at risk, i.e., classmates, teachers and staff, who had had close contact with the index case in a period starting from 48h before symptom onset of the symptomatic index case and, for asymptomatic cases, 48h before diagnosis or 48h after contact with a confirmed case, whichever occurred first. Mean attack rate was calculated as the mean value of all class-level transmissions.

A classroom cluster was defined as the presence of two or more positive students or teachers/staff members in the same class.

In this analysis we consider each single case or outbreak within a class as one statistical unit. For each class an index case was identified, i.e., the first case who tested positive (considering the date the swab was done). If more than one case in a class tested positive on the same day, the one with the earliest symptom onset was considered the index case. The same class can be included more than once because it may have been involved in more than one investigation during the study period.

When more than one class was included in a between-class transmission or there was a single index case for more than one class (usually, but not only, when the index case was a teacher), this was considered a school cluster.

If a classmate was already in isolation prior to symptom onset or swab positivity of the index case, due to contact with a positive person or re-entry from abroad, he/she was excluded from the denominator. Any student or staff who refused to perform a swab was excluded from the denominator. Cluster characteristics are reported; instead, information on the characteristics of cases diagnosed outside the province or that were identified during some experimental screening programme (2 classes) are missing.

Index cases were classified as student or teacher/staff and according to whether the case had a known contact outside the school setting (household, social activity, sport activity, not identified, and other/unspecified). The classes were classified according to the type of school (infant-toddler centre and preschool, primary school, lower secondary school, upper secondary school) and whether they were part of a multiclass cluster.

Delay in diagnosis of the index case, computed only for symptomatic cases, as the number of days between symptom onset and the date of swab positivity. Delay in contact tracing was computed as the time from swab positivity of the index case to the date on which the swab for (the majority of) classmates was scheduled.

Descriptive statistic was used to analyse and present the data. Data were analysed using Microsoft Excel and Stata v.13.1 (Statacorp, Tx).

### Ethical statement

The study was approved by the Area Vasta Emilia Nord Ethics Committee (no. 2020/0045199). The Ethics Committee authorised the use of subject data even in the absence of consent for people that could not be reached, if every reasonable effort had been made to contact that subject.

## Results

A total of 1213 student and 391 teacher/staff index cases were identified, corresponding to 1882 investigated classes, because some index cases, particularly teachers, had contacts in more than one class. A total of 43,792 swabs were requested for contact tracing in these classes; 43,214 (98.7%) were performed (table 1). The number of students and teachers/staff involved in contact tracing was about slightly less than half of the number of total resident students and teachers/staff in the province. However, this does not mean that epidemiological investigations involved this proportion of the school population, because some classes might have been involved in more than one investigation during the study period. Teacher index cases were more frequent in preschools and primary schools than in secondary schools.

**Table 1.**
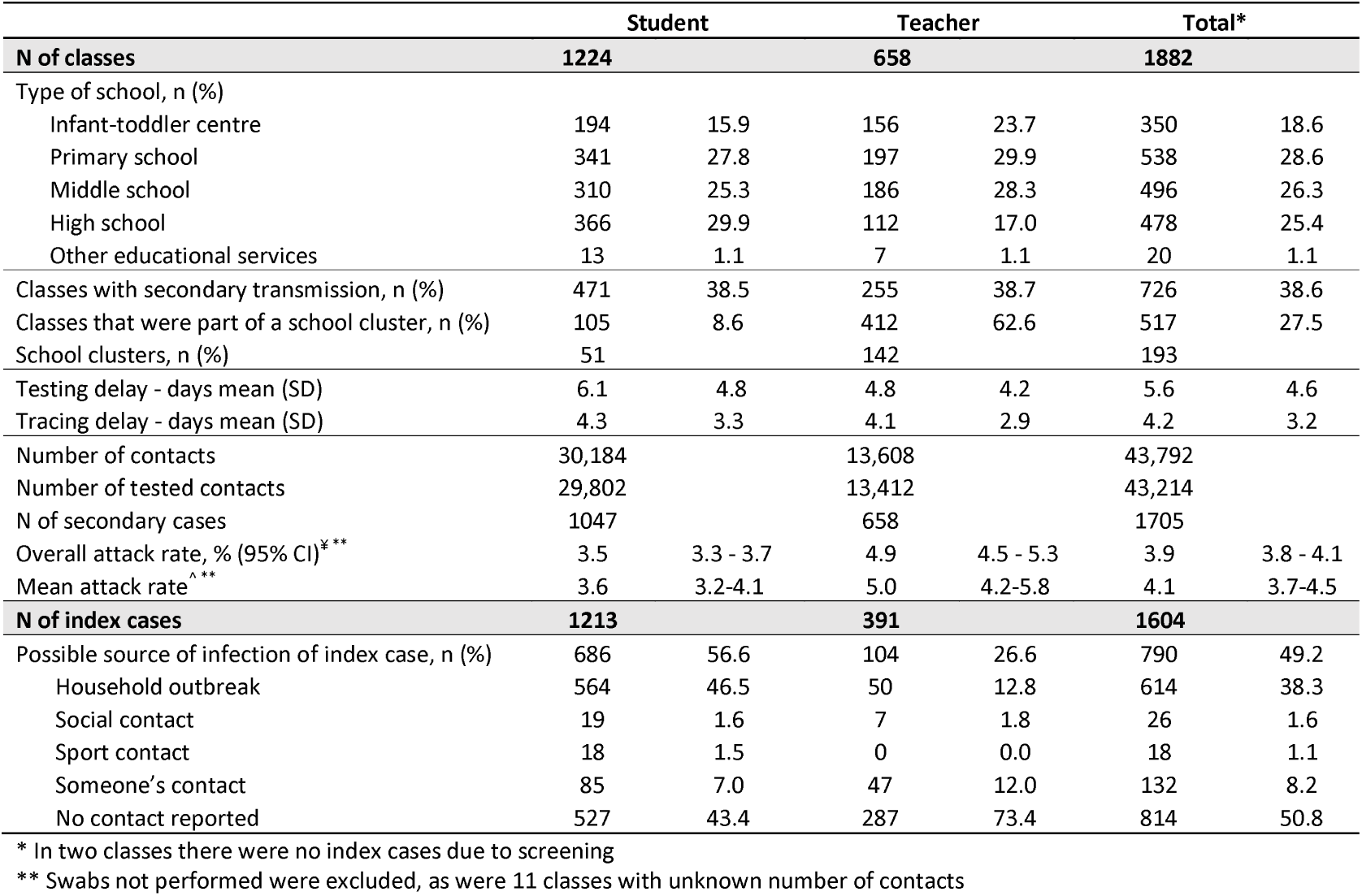
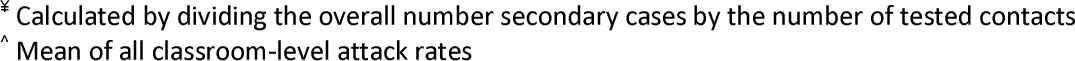
Characteristics of the field investigation in 1884* classes for which a school contact with a COVID-19 case was suspected, by type of index case. Reggio Emilia, September 2020-March 2021

|  | Student |  | Teacher |  | Total* |  |
| --- | --- | --- | --- | --- | --- | --- |
| **N of classes** | **1224** |  | **658** |  | **1882** |  |
| Type of school, n (%) |  |  |  |  |  |  |
| Infant-toddler centre | 194 | 15.9 | 156 | 23.7 | 350 | 18.6 |
| Primary school | 341 | 27.8 | 197 | 29.9 | 538 | 28.6 |
| Middle school | 310 | 25.3 | 186 | 28.3 | 496 | 26.3 |
| High school | 366 | 29.9 | 112 | 17.0 | 478 | 25.4 |
| Other educational services | 13 | 1.1 | 7 | 1.1 | 20 | 1.1 |
| Classes with secondary transmission, n (%) | 471 | 38.5 | 255 | 38.7 | 726 | 38.6 |
| Classes that were part of a school cluster, n (%) | 105 | 8.6 | 412 | 62.6 | 517 | 27.5 |
| School clusters, n (%) | 51 |  | 142 |  | 193 |  |
| Testing delay - days mean (SD) | 6.1 | 4.8 | 4.8 | 4.2 | 5.6 | 4.6 |
| Tracing delay - days mean (SD) | 4.3 | 3.3 | 4.1 | 2.9 | 4.2 | 3.2 |
| Number of contacts | 30,184 |  | 13,608 |  | 43,792 |  |
| Number of tested contacts | 29,802 |  | 13,412 |  | 43,214 |  |
| N of secondary cases | 1047 |  | 658 |  | 1705 |  |
| Overall attack rate, % (95% CI)<sup>***</sup> | 3.5 | 3.3 - 3.7 | 4.9 | 4.5 - 5.3 | 3.9 | 3.8 - 4.1 |
| Mean attack rate<sup>^**</sup> | 3.6 | 3.2-4.1 | 5.0 | 4.2-5.8 | 4.1 | 3.7-4.5 |
| **N of index cases** | **1213** |  | **391** |  | **1604** |  |
| Possible source of infection of index case, n (%) | 686 | 56.6 | 104 | 26.6 | 790 | 49.2 |
| Household outbreak | 564 | 46.5 | 50 | 12.8 | 614 | 38.3 |
| Social contact | 19 | 1.6 | 7 | 1.8 | 26 | 1.6 |
| Sport contact | 18 | 1.5 | 0 | 0.0 | 18 | 1.1 |
| Someone's contact | 85 | 7.0 | 47 | 12.0 | 132 | 8.2 |
| No contact reported | 527 | 43.4 | 287 | 73.4 | 814 | 50.8 |

\* In two classes there were no index cases due to screening
\*\* Swabs not performed were excluded, as were 11 classes with unknown number of contacts
\* Calculated by dividing the overall number secondary cases by the number of tested contacts
^ Mean of all classroom-level attack rates

Student index cases were identified in 56.5% of cases as a contact of other cases, the vast majority of which were within the same household (46.5%); a possible source of infection was identified for only 26.6% of teacher index cases (Table 1).

Secondary transmission occurred in less than 40% of classes, and the overall attack rate was 3.9% (95% CI 3.8-4.1%), 3.5% (95% CI 3.3-3.7%) for student index cases and 4.9% (95% CI 4.5-5.3%) for teacher index cases (Table 1). The attack rate over the study period was 4.8% in September and October, 2.6% in November, 3.9% in December and 4% from January to April (Table 2). The attack rate was similar in all types of schools (Table 3).

**Table 2.**
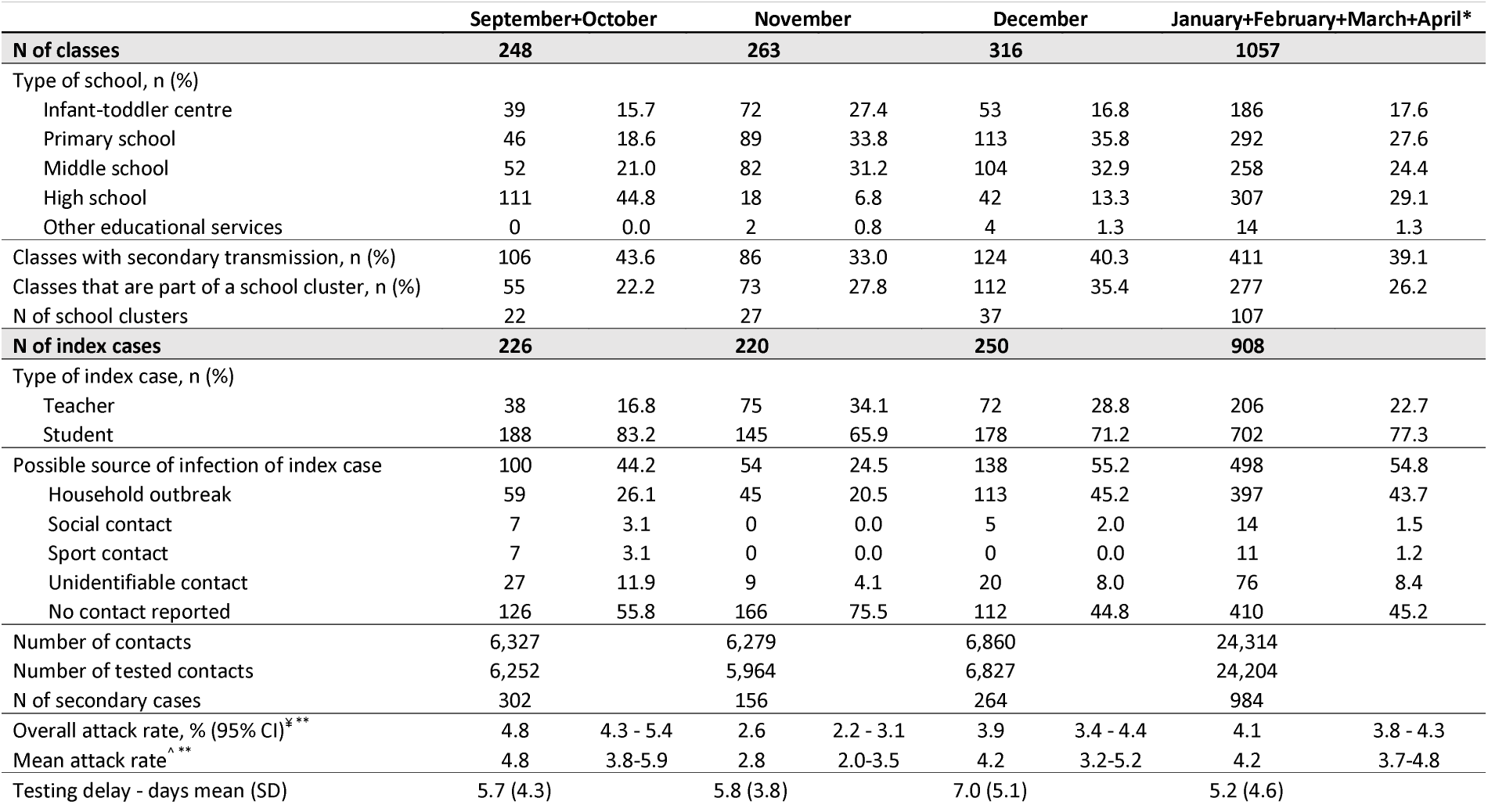
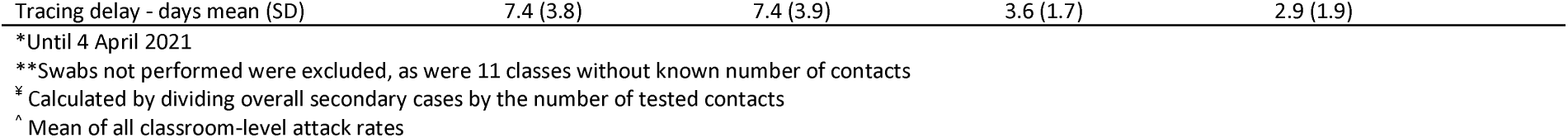
Characteristics of the field investigation in 1884 classes for which a school contact with a COVID-19 cases was suspected, by calendar period. Reggio Emilia, September 2020-March 2021

|  | September+October |  | November |  | December |  | January+February+March+April* |  |
| --- | --- | --- | --- | --- | --- | --- | --- | --- |
| **N of classes** | **248** |  | **263** |  | **316** |  | **1057** |  |
| Type of school, n (%) |  |  |  |  |  |  |  |  |
| Infant-toddler centre | 39 | 15.7 | 72 | 27.4 | 53 | 16.8 | 186 | 17.6 |
| Primary school | 46 | 18.6 | 89 | 33.8 | 113 | 35.8 | 292 | 27.6 |
| Middle school | 52 | 21.0 | 82 | 31.2 | 104 | 32.9 | 258 | 24.4 |
| High school | 111 | 44.8 | 18 | 6.8 | 42 | 13.3 | 307 | 29.1 |
| Other educational services | 0 | 0.0 | 2 | 0.8 | 4 | 1.3 | 14 | 1.3 |
| Classes with secondary transmission, n (%) | 106 | 43.6 | 86 | 33.0 | 124 | 40.3 | 411 | 39.1 |
| Classes that are part of a school cluster, n (%) | 55 | 22.2 | 73 | 27.8 | 112 | 35.4 | 277 | 26.2 |
| N of school clusters | 22 |  | 27 |  | 37 |  | 107 |  |
| **N of index cases** | **226** |  | **220** |  | **250** |  | **908** |  |
| Type of index case, n (%) |  |  |  |  |  |  |  |  |
| Teacher | 38 | 16.8 | 75 | 34.1 | 72 | 28.8 | 206 | 22.7 |
| Student | 188 | 83.2 | 145 | 65.9 | 178 | 71.2 | 702 | 77.3 |
| Possible source of infection of index case | 100 | 44.2 | 54 | 24.5 | 138 | 55.2 | 498 | 54.8 |
| Household outbreak | 59 | 26.1 | 45 | 20.5 | 113 | 45.2 | 397 | 43.7 |
| Social contact | 7 | 3.1 | 0 | 0.0 | 5 | 2.0 | 14 | 1.5 |
| Sport contact | 7 | 3.1 | 0 | 0.0 | 0 | 0.0 | 11 | 1.2 |
| Unidentifiable contact | 27 | 11.9 | 9 | 4.1 | 20 | 8.0 | 76 | 8.4 |
| No contact reported | 126 | 55.8 | 166 | 75.5 | 112 | 44.8 | 410 | 45.2 |
| Number of contacts | 6,327 |  | 6,279 |  | 6,860 |  | 24,314 |  |
| Number of tested contacts | 6,252 |  | 5,964 |  | 6,827 |  | 24,204 |  |
| N of secondary cases | 302 |  | 156 |  | 264 |  | 984 |  |
| Overall attack rate, % (95% CI)<sup>***</sup> | 4.8 | 4.3 - 5.4 | 2.6 | 2.2 - 3.1 | 3.9 | 3.4 - 4.4 | 4.1 | 3.8 - 4.3 |
| Mean attack rate<sup>^**</sup> | 4.8 | 3.8-5.9 | 2.8 | 2.0-3.5 | 4.2 | 3.2-5.2 | 4.2 | 3.7-4.8 |
| Testing delay - days mean (SD) | 5.7 (4.3) |  | 5.8 (3.8) |  | 7.0 (5.1) |  | 5.2 (4.6) |  |

|  |  |  |  |  |
| --- | --- | --- | --- | --- |
| Tracing delay - days mean (SD) | 7.4 (3.8) | 7.4 (3.9) | 3.6 (1.7) | 2.9 (1.9) |

\*Until 4 April 2021
\*\*Swabs not performed were excluded, as were 11 classes without known number of contacts
‡ Calculated by dividing overall secondary cases by the number of tested contacts
^ Mean of all classroom-level attack rates

**Table 3.** Characteristics of the field investigation in 1884 classes for which a school contact with a Covid-19 cases was suspected, by type of school. Reggio Emilia, September 2020-March 2021

|  | Infant-toddler centre |  | Primary school |  | Middle school |  | High school |  | Other educational services |  |
| --- | --- | --- | --- | --- | --- | --- | --- | --- | --- | --- |
| N of classes | 350 |  | 540 |  | 496 |  | 478 |  | 20 |  |
| Classes with secondary transmission | 131 | 37.4 | 216 | 40.0 | 173 | 34.9 | 202 | 42.3 | 5 | 25.0 |
| Classes that were part of a school cluster | 25 | 7.1 | 162 | 30.0 | 207 | 41.7 | 116 | 24.3 | 7 | 35.0 |
| N of school clusters | 10 |  | 70 |  | 68 |  | 43 |  | 2 |  |
| N of index cases | 338 |  | 466 |  | 370 |  | 415 |  | 15 |  |
| Type of index case, n (%) |  |  |  |  |  |  |  |  |  |  |
| Teacher | 144 | 42.6 | 126 | 27.0 | 66 | 17.8 | 51 | 12.3 | 4 | 26.7 |
| Student | 194 | 57.4 | 340 | 73.0 | 304 | 82.2 | 364 | 87.7 | 11 | 73.3 |
| Number of contacts | 7,767 |  | 11,709 |  | 12,106 |  | 11,936 |  | 306 |  |
| Number of tested contacts | 7,652 |  | 11,575 |  | 11,923 |  | 11,799 |  | 298 |  |
| N of secondary cases | 349 |  | 553 |  | 386 |  | 409 |  | 9 |  |
| Overall attack rate, % (95% CI)<sup>*,*</sup> | 4.6 | 4.1 - 5.1 | 4.8 | 4.4 - 5.2 | 3.2 | 2.9 - 3.6 | 3.5 | 3.1 - 3.8 | 3.0 | 1.4 - 5.7 |
| Mean attack rate<sup>^,*</sup> | 4.8 | 3.7-5.9 | 5.0 | 4.1-5.9 | 3.4 | 2.8-4.0 | 3.3 | 2.8-3.9 | 2.1 | 0.3-4.5 |
| Testing delay - days mean (SD) | 6.0 | 4.6 | 5.8 | 4.9 | 5.4 | 4.6 | 5.4 | 4.3 | 6.7 | 5.1 |
| Tracing delay - days mean (SD) | 4.4 | 3.5 | 4.1 | 3.1 | 4.1 | 3.3 | 4.4 | 3.0 | 3.3 | 2.3 |

\*Swabs not performed were excluded, as were 11 classes without known number of contacts
\* Calculated by dividing the overall number of secondary cases by the number of tested contacts
<sup>^</sup> Mean of all classroom-level attack rates

The mean delay in testing was 6 days in student index cases and 4.8 in teachers (Table 1); this did not change over the study period, except for a slight increase in December, and was similar across types of schools (Table 3). On the contrary, the delay in contact tracing decreased from December 2020 on, from about 7 days to 3 days (Table 2), not showing any differences between type of index case and type of school (Table 3).

## Discussion

We confirm a modest secondary attack rate in schools, as already observed in previous studies.^16-19^ Secondary cases occurred in about 40% of classes that were exposed to an infected classmate or teacher, and the overall attack rate was about 4%. Differently from our previous report^14^ and other studies reporting data on school cluster investigations,^19^ the attack rate was similar in preschools, primary schools and secondary schools. In fact, compared with the first two months of the study period,^14^ the attack rate increased in preschools from almost no transmission to 4%, while it remained stable in secondary schools. It must be considered that remote learning and periodic closures were implemented much more frequently in secondary schools, starting from 26 October 2020. Furthermore, an increase in the secondary case attack rate starting form late December/January was expected because the spread of the Alpha variant, which is more infectious than the previously circulating variants,^20^ started in that period in our region.^15^

Surprisingly, the difference in attack rate between classes where the index case was a student and those where the index case was a teacher was also small, despite the different kind of interactions and length of exposure of student-student contact and teacher-student contacts in the school setting. Unfortunately, we do not have any analytical information to define the quality and the duration of interactions between index cases and contacts.

The effect of changes in contact tracing strategies introduced mainly after the second wave of the pandemic, i.e., at the end of November 2020, was appreciable in the reduction in the delay in contact tracing and in the increase in the proportion of index cases for which a contact, particularly a contact in their household, was known. Indeed, a higher proportion of school investigations with a known link to household clusters was precisely the anticipated effect of introducing immediate testing of all asymptomatic contacts for all incident cases in the community, because this backward tracing made it possible to identify asymptomatic or paucisymptomatic students and to link them to their school contacts.

### Limitations and strengths

The main limitation of this study is that any observed association between changes in secondary attack rate and school or index case characteristics and especially actions implemented during the study period could be confounded by other factors that we could not measure. Among these, the most important was the spread of the Alpha variant, which probably started in our region around Christmas and which became the dominant circulating variant in February. Unfortunately, as very few cases were sequenced in Italy in that period,^15^ we cannot determine which clusters were due to the Alpha variant. Other variables that were missing or difficult to standardize were the duration and intensity of contacts between the index case and other classmates and the control measures each school independently put in place both before the occurrence of the case and during the outbreak.

The main strength of the study is its population-based nature; it included all cases for a which a school contact or exposure was identified. For an analysis of routine surveillance data, the completeness of field investigations is surprisingly high; only a few classes were not characterized, and very few classmates refused to be tested. Each outbreak in each school was investigated and re-checked for plausibility by the investigation team, thereby partially overcoming the shortcomings of cross-sectional, retrospective and self-reported studies.

## Conclusions

Overall, we confirmed a moderate risk of secondary transmission in the school setting (4.1% vs 3.2% in the previous study up to 15th October 2020), similar across all types of schools. The attack rate was also stable during the study period, despite the fact that the Alpha variant, which is much more infectious than the previously circulating variants, became predominant in the last included months. The promptness of contact tracing increased over the study period, reducing the time from index case identification to testing of contacts from 7 to 3 days, as well the ability to identify possible sources of infection in the index case.

## Data Availability

The data that support the findings of this study are available on request from the corresponding author.

## Acknowledgements

We thank all the Public Health Department officers and COVID-19 reference persons in the schools that collaborated in the investigations.

We thank Jacqueline M. Costa for the English language editing.

## Authors’ contributions

PGR, EL, MV and PP designed the study. EL, MC, SC, EB, DP, FV and EB contributed to the collection and management of the data. OD and LB analysed the data. OD and PGR interpreted the data and wrote the manuscript. AMU contributed to data analysis and interpretation of results. All authors revised the manuscript and approved the final version.

## What is already known on this topic

- Closure of educational institutions was one of the preventive measures considered and often adopted during the pandemic.
- The role of school contacts in the spread of the virus as well the effectiveness of school closures in controlling the epidemic is still debated.
- Potential effect of the more prompt (backward) contact tracing on secondary transmission in educational setting has not been evaluated.

## What this study adds

- Risk of transmission in educational setting is moderate with stabile attack rates over time and by type of school.
- Secondary attack rate is slightly higher if index case is a teacher.
- The tracing delay has halved after introducing the backward contact tracing, while ability to identify possible sources of infection in the index case increased.

## References

1. Dong E, Du H, Gardner L. An interactive web-based dashboard to track COVID-19 in real time. Lancet Infect Dis. 2020;20(5):533–534. doi: 10.1016/S1473-3099(20)30120-1.

2. Viner RM, Mytton OT, Bonnel C, Mellendez-Torres JG, Ward J, Hudson L, et al. Susceptibility to SARS-CoV-2 Infection Among Children and Adolescents Compared With Adults. A Systematic Review and Meta-analysis. JAMA Pediatr. 2021;175(2):143-156. Doi: 10.1001/jamapediatrics.2020.4573

3. Gaythorpe, KAM, Bhatia S, Mangal T, et al. Children’s role in the COVID-19 pandemic: a systematic review of early surveillance data on susceptibility, severity, and transmissibility. 2021;11(1):13903. doi: 10.1038/s41598-021-92500-9.

4. Haug N, Geyrhofer L, Londei A, Dervic E, Desvars-Larrive A, Loreto V, Pinior B, Thurner S, Klimek P. Ranking the effectiveness of worldwide COVID-19 government interventions. Nat Hum Behav. 2020;4(12):1303–1312. doi: 10.1038/s41562-020-01009-0.

5. Head JR, Andrejko K, Cheng Q, et al. The effect of school closures and reopening strategies on COVID-19 infection dynamics in the San Francisco Bay Area: a cross-sectional survey and modeling analysis. medRxiv [Preprint]. 2020. doi: 10.1101/2020.08.06.20169797.

6. Keeling MJ, Tildesley MJ, Atkins BD, et al. The impact of school reopening on the spread of COVID-19 in England. Philos Trans R Soc Lond B Biol Sci. 2021;376(1829):20200261 doi: 10.1098/rstb.2020.0261.

7. Landeros A, Ji X, Lange K, et al. An examination of school reopening strategies during the SARS-CoV-2 pandemic. PLoS One. 2021;16(5):e0251242. doi: 10.1371/journal.pone.0251242.

8. Walsh S, Chowdhury A, Russell S, et al. Do school closures reduce community transmission of COVID-19? A systematic review of observational studies MedRxiv. 2021. doi: https://doi.org/10.1101/2021.01.02.21249146.

9. Courtemanche C, Garuccio J, Le A, et al. Strong Social Distancing Measures In The United States Reduced The COVID-19 Growth Rate. Health Aff (Millwood) 2020; 39: 1237–46. doi: 10.1377/hlthaff.2020.00608.

10. Hsiang S, Allen D, Annan-Phan S, et al. The effect of large-scale anti-contagion policies on the COVID-19 pandemic. Nature 2020; 584: 262–7. doi: 10.1038/s41586-020-2404-8.

11. Ertem Z, Schechter-Perkins E, Oster E, et al. The Impact of School Opening Model on SARS-CoV-2 Community Incidence and Mortality: A Nationwide Cohort Study. Res Sq [Preprint]. 2021 Jul 15:rs.3.rs-712725. doi: 10.21203/rs.3.rs-712725/v1.

12. Forbes H, Morton CE, Bacon S, et al. Association between living with children and outcomes from covid-19: OpenSAFELY cohort study of 12 million adults in England. BMJ. 2021;372:n628. doi: 10.1136/bmj.n628.

13. Marziano V, Guzzetta G, Rondinone BM, et al. Retrospective analysis of the Italian exit strategy from COVID-19 lockdown. Proc Natl Acad Sci U S A. 2021;118(4):e2019617118. doi: 10.1073/pnas.2019617118.

14. Larosa E, Djuric O, Cassinadri M, et al. Secondary transmission of COVID-19 in preschool and school settings in northern Italy after their reopening in September 2020: a population-based study. Euro Surveill. 2020;25(49):2001911. doi: 10.2807/1560-7917.ES.2020.25.49.2001911.

15. Istituto Superiore di Sanità. Prevalenza e distribuzione delle varianti di SARS-CoV-2 di interesse per la sanità pubblica in Italia. Rapporto n. 5 del 23 luglio 2021. July 2021. [Italian] Available at: https://www.epicentro.iss.it/coronavirus/pdf/sars-cov-2-monitoraggio-varianti-rapporti-periodici-23-luglio-2021.pdf

16. Ehrhardt J, Ekinci A, Krehl H, Meincke M, Finci I, Klein J, et al. Transmission of SARS-CoV-2 in children aged 0 to 19 years in childcare facilities and schools after their reopening in May 2020, Baden-Württemberg, Germany. Euro Surveill. 2020;25(36):2001587. doi: 10.2807/1560-7917.ES.2020.25.36.2001587

17. Macartney K, Quinn HE, Pillsbury AJ, Koirala A, Deng L, Winkler N, et al. Transmission of SARS-CoV-2 in Australian educational settings: a prospective cohort study. Lancet Child Adolesc Health. 2020;4(11):807–816. doi: 10.1016/S2352-4642(20)30251-0

18. Yoon Y, Kim KR, Park H, et al. Stepwise school opening online and off-line and an impact on the epidemiology of COVID-19 in the pediatric population. MedRxiv.2020; (Preprint). doi: 10.1101/2020.08.03.20165589.

19. Aiano F, Mensah AA, McOwat K, Obi C, Vusirikala A, Powell AA, Flood J, Bosowski J, Letley L, Jones S, Amin-Chowdhury Z, Lacy J, Hayden I, Ismail SA, Ramsay ME, Ladhani SN, Saliba V. COVID-19 outbreaks following full reopening of primary and secondary schools in England: Cross-sectional national surveillance, November 2020. Lancet Reg Health Eur. 2021 Jul;6:100120. doi: 10.1016/j.lanepe.2021.100120. Epub 2021 May 19.

20. Davies NG, Abbott S, Barnard RC, et al. Estimated transmissibility and impact of SARS-CoV-2 lineage B.1.1.7 in England. Science. 2021;372(6538):eabg3055. doi: 10.1126/science.abg3055.

